# Efficient Surveillance and Temporal Calibration of Disease Response

**DOI:** 10.1101/2021.01.13.21249781

**Authors:** Kamran Najeebullah, Jessica Liebig, Jonathan Darbro, Raja Jurdak, Dean Paini

## Abstract

**Background:** Disease surveillance and response are critical components of epidemic preparedness. The disease response, in most cases, is a set of reactive measures that follow the outcomes of the disease surveillance. Hence, efficient surveillance is a prerequisite for an effective response.

**Methodology/Principal Findings:** We introduce the principle of epidemiological soundness and utilize it to construct a disease spread network. The network implicitly quantifies the fertility (whether a case leads to secondary cases) and reproduction (number of secondary cases per infectious case) of the cases as well as the size and generations (of the infection chain) of the outbreaks. We empirically confirm that high morbidity relates positively with delay in disease response. Moreover, we identify what constitutes efficient surveillance by applying various thresholds of disease response delay to the network and report their impact on case fertility, reproduction, number of generations and ultimately, outbreak size. Lastly, we identify the components of the disease surveillance system that can be calibrated to achieve the identified efficiency thresholds.

**Conclusion:** We identify practically achievable, efficient surveillance thresholds (on temporal scale) that lead to an effective response and identify how they can be satisfied. Our approach can be utilized to provide guidelines on spatially and demographically targeted resource allocation for public awareness campaigns as well as to improve diagnostic abilities and turn-around times for the doctors and laboratories involved.

**Author Summary:** Efficient surveillance and effective response capabilities are pivotal to the prevention and control of the infectious diseases. The disease response is a set of reactive actions that follow the outcomes of the disease surveillance. Ergo, efficient surveillance is a perquisite for the deployment of an effective response. The quantification of the efficiency of a disease surveillance system largely depends on the epidemiological characteristics of the disease. In this paper, we introduce an approach that builds on these characteristics and measures the performance of a disease surveillance system through its impact on the incidence of the disease. Using this approach, we obtain quantitative (on a temporal scale) efficient surveillance thresholds, which if followed by a timely response, lead to a considerable reduction in the disease incidence. Furthermore, we show that these thresholds are practically achievable by identifying the obstacles that lead to less than efficient surveillance outcomes. Our approach can be applied to obtain guidelines on spatially, temporally and demographically targeted resource allocations for public awareness campaigns as well to improve diagnostic ability and turn-around times in treating doctors and pathology labs.

## Introduction

Sporadic outbursts of emerging and re-emerging infectious diseases are on the rise, carrying catastrophic consequences for health and livelihoods of people in poor and wealthy parts of the world alike [1–3]. Fueled by the effects of globalization, the growing mobility of human populations and constant urbanization, epidemics can reach pandemic scale in a matter of days [4]. A warming and unstable climate, increasing population density and frequent interactions between humans and wild animals are likely to further amplify the risk of emerging diseases [3,4], posing an imminent threat to global health security.

Epidemic preparedness is an indispensable element of health security in mitigating the threat of infectious diseases. Efficient surveillance and effective response capabilities are pivotal for epidemic preparedness [5]. A disease surveillance system is a tool to detect, confirm and report the occurrence of a disease in a population. Disease response, on the other hand, is a set of reactive (to the outcome of surveillance) actions to reduce morbidity and consequent mortality [6]. Disease response is not an exact science and the actions to control an infectious disease may differ due to variations in factors including seasonality, climate, individual susceptibility, community values, and characteristics of the pathogen itself. Regardless of the context, experts agree that minimizing response time is critical for epidemic control [7–9]. Disease response inherently builds on the outcomes of disease surveillance and cannot precede it temporally. Thus implying, efficient surveillance leads to effective response.

In this paper we introduce a methodology that quantifies the efficiency of a disease surveillance system. To this end, we construct a *disease spread network* to identify disease pathways by defining parent-child relationships between the individual cases. We introduce the *principle of epidemiological soundness* that utilizes the disease occurrence data in combination with the meteorological and human population mobility data to estimate these relations. Unlike prior studies where the timeliness of the disease surveillance is measured by comparing the average delays in case notification to a predefined standard [10,11], our methodology quantifies the efficiency by measuring the case fertility (whether a case leads to secondary cases) and reproduction (number of secondary cases per infectious case) as well as the outbreak size and generations (of the infection chain). While the traditional approach is important to identify the efficiency gaps, our approach goes a step further by measuring the impact of these gaps on the incidence of the disease. The construction of the disease spread network allows us to empirically confirm that high morbidity relates positively with delay in disease response. Furthermore, we test varying thresholds for what may represent efficient surveillance and gauge their impact on various parameters associated with the disease occurrence. Lastly, we conduct a spatio-temporal analysis of the delays involved at the various stages of the disease surveillance, identify efficiency bottlenecks, and provide insights on how they may be prevented. Our results can be utilized to provide guidelines on spatially and demographically targeted resource allocation for public awareness campaigns as well as to improve diagnostic abilities and turn-around times for the doctors and laboratories involved. The efficient surveillance thresholds obtained in this study can be utilized to define a case ranking system that can help prioritize testing of the cases that are more likely to lead to a larger outbreaks.

## Methods and Materials

### Notions

We choose dengue as the infectious disease to implement our methodology. Our choice is motivated by the availability of data. Dengue is transmitted when an infectious female vector, primarily *Aedes aegypti*, bites a susceptible person, formally known as *time of exposure*. Symptoms of the infection start to appear within 3 to 10 days [12], a period of time known as the *intrinsic incubation period* (IIP). The *latent period* (LP) ends up to 2 days prior to the onset of symptoms [13] and can only be observed through a lab test. A person is capable of providing an infectious blood meal after the end of the latent period, and retains this capability for up to 7 days after the symptom onset [14]. This period is referred to as the *infectious period*. A susceptible mosquito undergoes an *extrinsic incubation period* (EIP) after having an infectious blood meal. The duration of the EIP is influenced by a range of factors including the ambient temperature, and has been reported to be as short as 3 days [12]. In unfavourable conditions (e.g., when temperatures drop below 20° Celsius), the EIP is estimated to be longer than the typical mosquito’s lifetime [12]. In order to facilitate our analysis, we introduce the notion of *transmission period*. The transmission period begins at the end of the EIP and persists until the death of the infectious mosquito. Since an infectious person may infect multiple mosquitoes, the transmission period for a particular dengue case is an aggregation of transmission periods of all the infected mosquitoes. Starting with the earliest concluding EIP and ending with the death of the last infected mosquito. See Figure 1 for a visual depiction of the dengue infection timeline.

**Figure 1.**
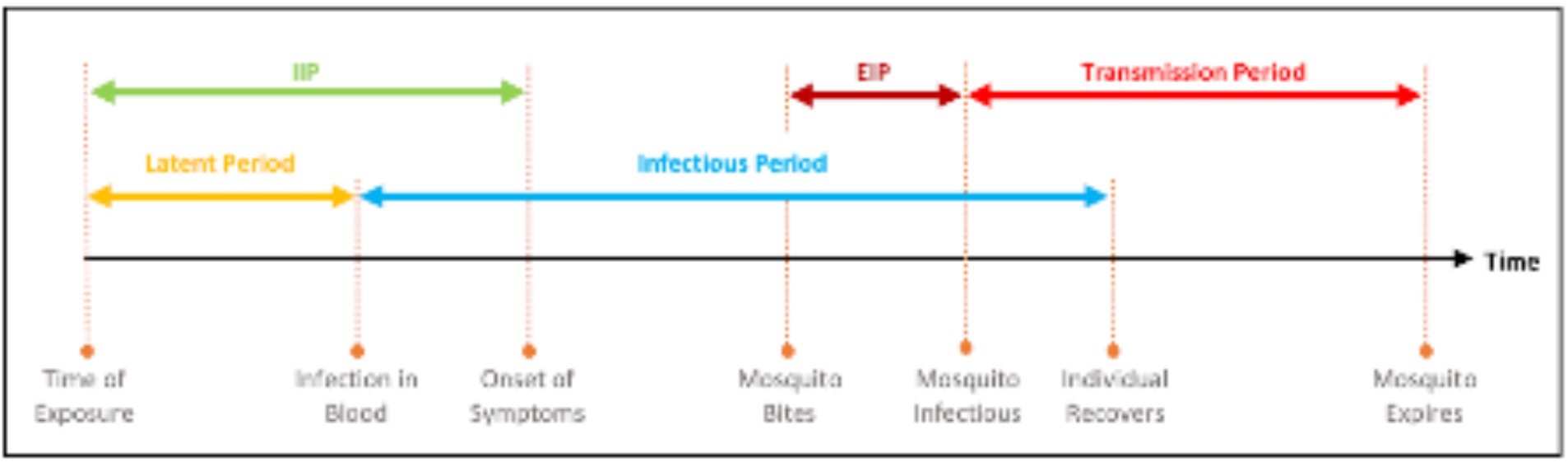
A visual depiction of the dengue infection timeline. Arrows marked IIP and EIP represent the Intrinsic and Extrinsic Incubation Period, respectively.

### Dengue in Australia

To evaluate our methodology, we take the reported occurrence of dengue in Australia, between 2002 and 2018, as a case study. Clearly, the transmission of dengue is subject to the presence of the vector. In Australia, the vector presence is limited to parts of the state of Queensland. The primary vector, Aedes aegypti, is prevalent in northern coastal communities and has also been recorded in several areas of central and southern Queensland [15,16]. The presence of *Aedes albopictus*, a secondary vector capable of transmitting dengue infection, has also been recorded in the northern parts (Torres Strait Islands) of the state [17].

Queensland Health has a passive surveillance system in place for identifying dengue cases. The system relies on general practitioners, emergency departments and laboratories notifying Queensland Health of suspected and laboratory confirmed cases. The treating doctors are required to report a case upon clinical suspicion under the provision of the Public Health Act 2005 [18]. Since dengue is not endemic in Australia, particular attention is paid to people who exhibit dengue symptoms and have recently travelled to an endemic country. Queensland Health defines a dengue outbreak as at least one locally acquired case [16]. All laboratory-confirmed and suspected cases are notified to the local Public Health Unit (PHU). Upon notification the PHU communicates with the patient as well as the treating doctor and laboratory to collect case details. This includes the spatio-temporal aspects of the case(see Data Collection section for details).

Vector elimination is the first line of defence for Queensland Health to prevent the wider spread of dengue. According to the Queensland Dengue Management Plan [16], when a dengue case is notified it is referred to a medical entomologist by the concerned PHU. If the reported case is in a region with known vector presence, the medical entomologist instigates appropriate vector control measures. The measures consist of indoor residual spraying and treating water containers using insect growth regulators or pesticides at the addresses identified through the travel and contact history of the patient. The aim is to eradicate the virus by eliminating the vector within a radius of about 200 metres (the estimated distance a vector can travel over its lifetime) [19]. Queensland Health works collaboratively with local governments and dedicated Dengue Action Response Teams to manage the efforts.

Since vector control in Queensland is a reactive exercise, it is more effective when conducted closer in time to the start of the infectious period. Theoretically, if conducted within the EIP (the beginning of EIP and infectious period can coincide as a susceptible mosquito may bite the infectious person on the first day of infectiousness), the efforts can potentially prevent any subsequent local transmission.

However, prompt reporting of a case may not always be possible due to various factors, including awareness and willingness of a patient to seek medical advice, training and diagnostic ability of the treating doctor, capability and capacity of the testing laboratory and efficiency and complexity of the disease surveillance mechanism. We call the time elapsed (in days) since a case has become infectious until it has been notified to Queensland Health the *notification delay*.

### Data Collection

Dengue is a notifiable disease in Australia under the Public Health Act 2005 and laboratory confirmed cases are required to be notified to health departments within each state and territory [18]. Since the impact of disease response can only be measured in the regions with local transmission, we limit the scope of our analysis to these regions. Data on the occurrence of dengue was obtained from the Communicable Disease Branch (CDB) of Queensland Health for the period of October 2001 to September 2018. During this period 5,272 cases were reported, of which 2,808 (53%) were acquired locally. The data records cases on an individual level and covers their spatial and temporal aspects. Spatial information includes the place of residence and the place of acquisition on locality/suburb and postcode levels, respectively. For cases acquired overseas the highest resolution for the place of acquisition is the country. Temporal information consists of the date of symptom onset, specimen collection, diagnosis and notification.

Traditionally, temporal analysis of the dengue cases is performed with an observation window set for each calendar year, where cases are assigned to a window with respect to their date of symptom onset. Since the majority of locally acquired dengue cases in Queensland occur between October and May, we move the observation window to align it with this seasonal fluctuation. For each year, our observation window starts at the beginning of September of the previous year and concludes at the end of August of the current year. This shift allows us to incorporate the outbreaks that span across calendar years (see Table 1 for a year-wise breakdown of the occurrence). For the ease of analysis, spatial statistics are aggregated by Statistical Area Level 4 (SA4). The SA4s are sub-State regions defined by the Australian Statistical Geography Standard (ASGS) with a population range between 100,000 and 500,000 individuals. Queensland has 19 SA4 regions in total, of which 7, namely Cairns, Darling Downs, Fitzroy, Mackay, Queensland-Outback, Townsville and Wide Bay, have recorded the presence of the vector.

**Table 1.**
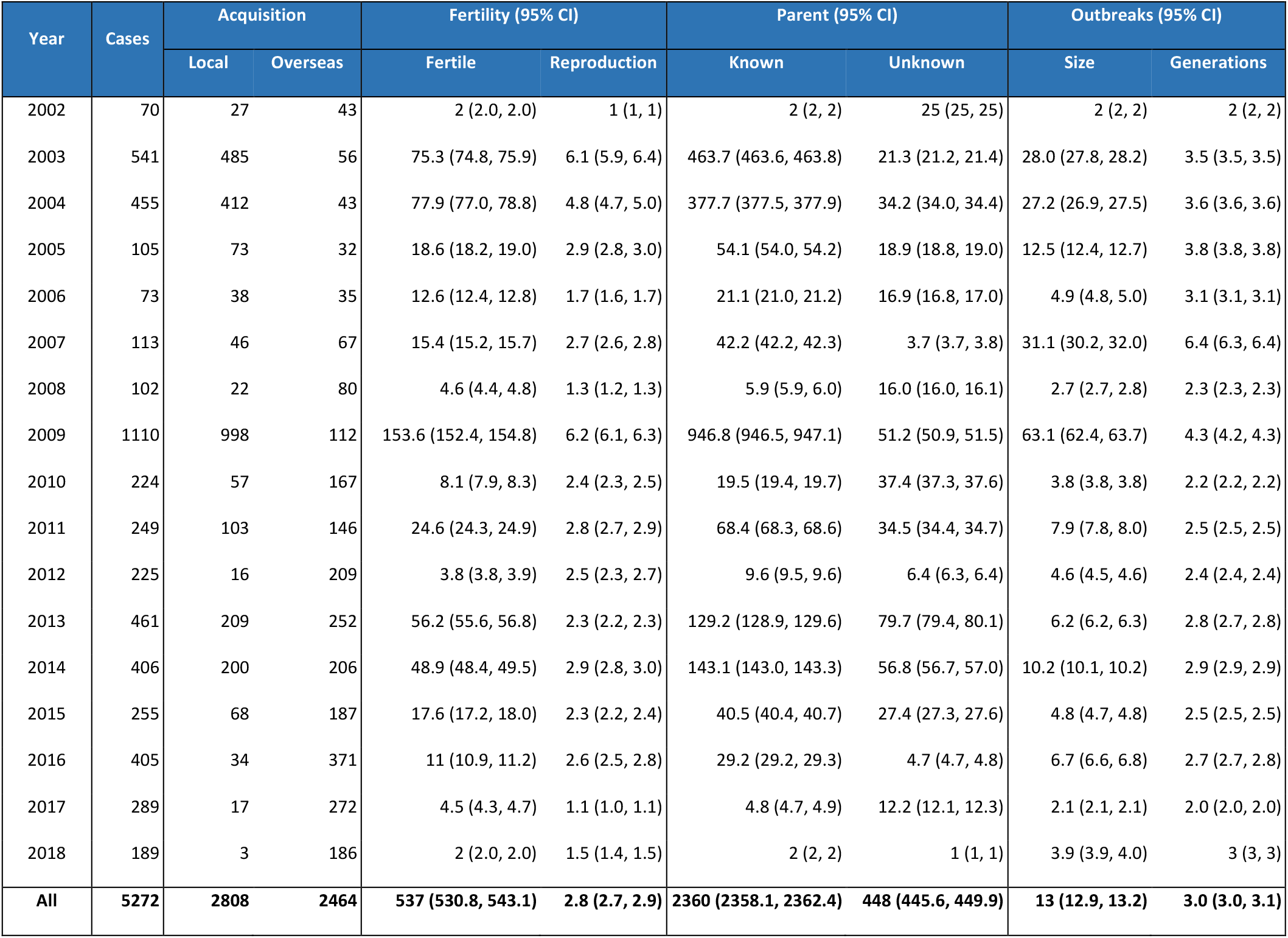
Classification of dengue occurrence in Queensland, Australia. The Fertility, Parent and Outbreak columns present statistics in the context of the disease spread network. A case is fertile if it has resulted in at least one secondary case and the reproduction indicates the number of secondary cases caused by a case. The parent of a locally acquired case is unknown if no suitable parent case was found during the construction of the disease spread network and known otherwise.

| Year | Cases | Acquisition |  | Fertility (95% CI) |  | Parent (95% CI) |  | Outbreaks (95% CI) |  |
| --- | --- | --- | --- | --- | --- | --- | --- | --- | --- |
|  |  | Local | Overseas | Fertile | Reproduction | Known | Unknown | Size | Generations |
| 2002 | 70 | 27 | 43 | 2 (2.0, 2.0) | 1 (1, 1) | 2 (2, 2) | 25 (25, 25) | 2 (2, 2) | 2 (2, 2) |
| 2003 | 541 | 485 | 56 | 75.3 (74.8, 75.9) | 6.1 (5.9, 6.4) | 463.7 (463.6, 463.8) | 21.3 (21.2, 21.4) | 28.0 (27.8, 28.2) | 3.5 (3.5, 3.5) |
| 2004 | 455 | 412 | 43 | 77.9 (77.0, 78.8) | 4.8 (4.7, 5.0) | 377.7 (377.5, 377.9) | 34.2 (34.0, 34.4) | 27.2 (26.9, 27.5) | 3.6 (3.6, 3.6) |
| 2005 | 105 | 73 | 32 | 18.6 (18.2, 19.0) | 2.9 (2.8, 3.0) | 54.1 (54.0, 54.2) | 18.9 (18.8, 19.0) | 12.5 (12.4, 12.7) | 3.8 (3.8, 3.8) |
| 2006 | 73 | 38 | 35 | 12.6 (12.4, 12.8) | 1.7 (1.6, 1.7) | 21.1 (21.0, 21.2) | 16.9 (16.8, 17.0) | 4.9 (4.8, 5.0) | 3.1 (3.1, 3.1) |
| 2007 | 113 | 46 | 67 | 15.4 (15.2, 15.7) | 2.7 (2.6, 2.8) | 42.2 (42.2, 42.3) | 3.7 (3.7, 3.8) | 31.1 (30.2, 32.0) | 6.4 (6.3, 6.4) |
| 2008 | 102 | 22 | 80 | 4.6 (4.4, 4.8) | 1.3 (1.2, 1.3) | 5.9 (5.9, 6.0) | 16.0 (16.0, 16.1) | 2.7 (2.7, 2.8) | 2.3 (2.3, 2.3) |
| 2009 | 1110 | 998 | 112 | 153.6 (152.4, 154.8) | 6.2 (6.1, 6.3) | 946.8 (946.5, 947.1) | 51.2 (50.9, 51.5) | 63.1 (62.4, 63.7) | 4.3 (4.2, 4.3) |
| 2010 | 224 | 57 | 167 | 8.1 (7.9, 8.3) | 2.4 (2.3, 2.5) | 19.5 (19.4, 19.7) | 37.4 (37.3, 37.6) | 3.8 (3.8, 3.8) | 2.2 (2.2, 2.2) |
| 2011 | 249 | 103 | 146 | 24.6 (24.3, 24.9) | 2.8 (2.7, 2.9) | 68.4 (68.3, 68.6) | 34.5 (34.4, 34.7) | 7.9 (7.8, 8.0) | 2.5 (2.5, 2.5) |
| 2012 | 225 | 16 | 209 | 3.8 (3.8, 3.9) | 2.5 (2.3, 2.7) | 9.6 (9.5, 9.6) | 6.4 (6.3, 6.4) | 4.6 (4.5, 4.6) | 2.4 (2.4, 2.4) |
| 2013 | 461 | 209 | 252 | 56.2 (55.6, 56.8) | 2.3 (2.2, 2.3) | 129.2 (128.9, 129.6) | 79.7 (79.4, 80.1) | 6.2 (6.2, 6.3) | 2.8 (2.7, 2.8) |
| 2014 | 406 | 200 | 206 | 48.9 (48.4, 49.5) | 2.9 (2.8, 3.0) | 143.1 (143.0, 143.3) | 56.8 (56.7, 57.0) | 10.2 (10.1, 10.2) | 2.9 (2.9, 2.9) |
| 2015 | 255 | 68 | 187 | 17.6 (17.2, 18.0) | 2.3 (2.2, 2.4) | 40.5 (40.4, 40.7) | 27.4 (27.3, 27.6) | 4.8 (4.7, 4.8) | 2.5 (2.5, 2.5) |
| 2016 | 405 | 34 | 371 | 11 (10.9, 11.2) | 2.6 (2.5, 2.8) | 29.2 (29.2, 29.3) | 4.7 (4.7, 4.8) | 6.7 (6.6, 6.8) | 2.7 (2.7, 2.8) |
| 2017 | 289 | 17 | 272 | 4.5 (4.3, 4.7) | 1.1 (1.0, 1.1) | 4.8 (4.7, 4.9) | 12.2 (12.1, 12.3) | 2.1 (2.1, 2.1) | 2.0 (2.0, 2.0) |
| 2018 | 189 | 3 | 186 | 2 (2.0, 2.0) | 1.5 (1.4, 1.5) | 2 (2, 2) | 1 (1, 1) | 3.9 (3.9, 4.0) | 3 (3, 3) |
| **All** | **5272** | **2808** | **2464** | **537 (530.8, 543.1)** | **2.8 (2.7, 2.9)** | **2360 (2358.1, 2362.4)** | **448 (445.6, 449.9)** | **13 (12.9, 13.2)** | **3.0 (3.0, 3.1)** |

In order to infer relationships between the cases, the occurrence data is utilized in combination with weather and population mobility data. Daily weather data includes minimum and maximum temperatures for the period of October 2001 to September 2018, recorded at 216 weather stations across Queensland and was obtained from the Australian Bureau of Meteorology. For human mobility data, we constructed yearly origin-destination matrices for the studied period from National Visitor Surveys (NVS), International Visitor Surveys (IVS) and Twitter data. The surveys of national and international visitors were conducted by Tourism Research Australia and record the different SA4 regions that individuals visited. For national visitors the SA4 region of residence is also recorded. NVS data is available between 1998 and 2015, IVS data is available between 2005 and 2015. The Twitter data supplements the IVS and NVS datasets as a source of intra-region mobility. It was collected in 2015 and contains 925,945 trips from 79,271 individuals [20]. For the missing years, we apply an auto-regressive moving average model [21], a common method for forecasting time-series to estimate movement between regions.

### Disease Spread Network

Utilizing the dengue occurrence data along with the population mobility and meteorology datasets, we construct a probable *disease spread network* (DSN) that depicts the spatio-temporal spread of dengue in Queensland. The DSN is a directed acyclic graph, where nodes represent dengue cases and arcs represent probable relationships between them. The construction of the network is a two stage process. In the first stage, we draw arcs on the principle of epidemiological soundness and in the second stage, we prune the network using the Latent Influence Point Process model (LIPP) [22] to identify the most likely transmission paths.

The epidemiological soundness of an arc depends on the spatio-temporal and clinical characteristics of the cases that are being considered to be connected by the arc. More specifically, in order for two cases *p* and *c* to be connected via an arc (*p* → *c*), they must fulfill the following criteria;

- the exposure period of c must overlap with the transmission period of p,
- the exposure period of c must overlap with the transmission period of p,
- the region of acquisition of c must coincide with region of residence of p (applied on SA4 level),
- p and c must not be infected by different serotypes of dengue.

We describe the relationship between *p* and *c* as that of a parent and child, where *p* is the parent case and *c* is the child case. To fulfil the first criterion, we estimate the IIP and minimum EIP for each case using a Gamma and log-normal *time-to-event model* respectively, where the latter incorporates temperature sensitivity through a co-variate [12].

The LP is estimated by decreasing the estimated IIP value by 2 [13]. The remaining criteria can be tested using the information provided in the dengue occurrence dataset. The IIP is randomly drawn from a log-normal distribution and its randomness may impact the structure of the network. To account for this variability in our analysis we repeat the network construction process 100 times and present disagreement of measures across various instances through a 95% confidence interval wherever applicable. The EIP on the other hand, always yields the same value for a given temperature.

At the end of the first stage of network construction we obtain a probable DSN that logically aligns with the epidemiological characteristics of dengue. Yet it is not an acceptable representation of disease spread. In nature, we expect each dengue case to have exactly one parent case. However, the above criteria do not prevent a node from having multiple in-coming arcs, implying a practical impossibility that it has been caused by more than one case. The second stage of the network construction is required to address this problem.

In the second stage, we use the LIPP model to prune the excess arcs. The LIPP model estimates pair-wise causation probabilities for dengue cases using a multivariate *Hawkes process* [23]. It models disease spread across a heterogeneous social system by incorporating three major counterbalancing factors: (i) exogenous influence covering environmental heterogeneity, (ii) endogenous influence attributed to macro level interactions between meta-populations, and (iii) a time decay effect. The estimates are computed using the disease occurrence and population mobility data. We use the estimates generated by the LIPP model to break ties between multiple parents of a child case resulting in the most likely parent-child arc being preserved.

In the context of the disease spread network, we define a case to be fertile if the node representing the case has at least one out-going arc and infertile otherwise. Similarly the reproduction of a case denotes the number of out-going arcs of the node representing the case.

## Results

### Case Fertility and Response Delay

Of the 5,272 reported cases, 537 (10%) were estimated to be fertile. Moreover, of the 2,808 locally acquired case, a parent was found for 2,360 (84%) while no suitable candidates were found for 448 (16%) (see Table 1 for details on fertility and parent-child relationship estimates). The years with the highest number of fertile cases mostly correspond to years with most number of reported cases. However, in terms of proportion of reported cases, 2005 was estimated to have the highest proportion of fertile cases (18%) followed by 2006 and 2004 with 17% fertile cases each. In terms of region-wise distribution, Cairns was estimated to have the highest number of fertile cases (350) followed by Townsville (112) and Queensland-Outback (44). Proportionally, Townsville led Cairns with 22% fertile cases compared to 15%, with Queensland-Outback (10%) at the third spot.

We observed a positive relation between the fertile cases and the length of transmission period. Recall that the response delay of a case is the sum of the minimum EIP and the transmission period associated with the case. Hence a longer transmission period implies a longer response delay. Our estimation showed that over 75% of the 537 fertile cases had a transmission period of 5 or more days. Moreover, half of the fertile cases had a transmission period of at least 8 days (of which 50% had a transmission period of length between 14 and 28 days). On the other hand, only 5% of the fertile cases had a transmission period of 2 days or less. These observation indicate that timely notification and rapid response are critical in prevention of secondary cases.

A year-wise breakdown of the fertile cases (see Figure 2a) presented a similar pattern. With the exception of 2017, over half of the fertile cases reported each year had an estimated transmission period of at least 5 days. The same was the case for over 75% of the fertile cases reported in 10 out of the 16 years considered. Furthermore, more than half of the fertile cases reported in 8 years had a transmission period of 10 days or longer. Spatially (Figure 2b), fertile cases in Townsville were estimated to have the longest transmission periods on average, where 75% and 50% fertile cases had a transmission period of at least 7 and 11 days, respectively. Collectively, each region had at least 50% fertile cases with a transmission period in excess of 5 days. In fact, in Cairns and Queensland- Outback, half of the fertile cases had a transmission period of at least 7 and 8 days, respectively.

**Figure 2.**
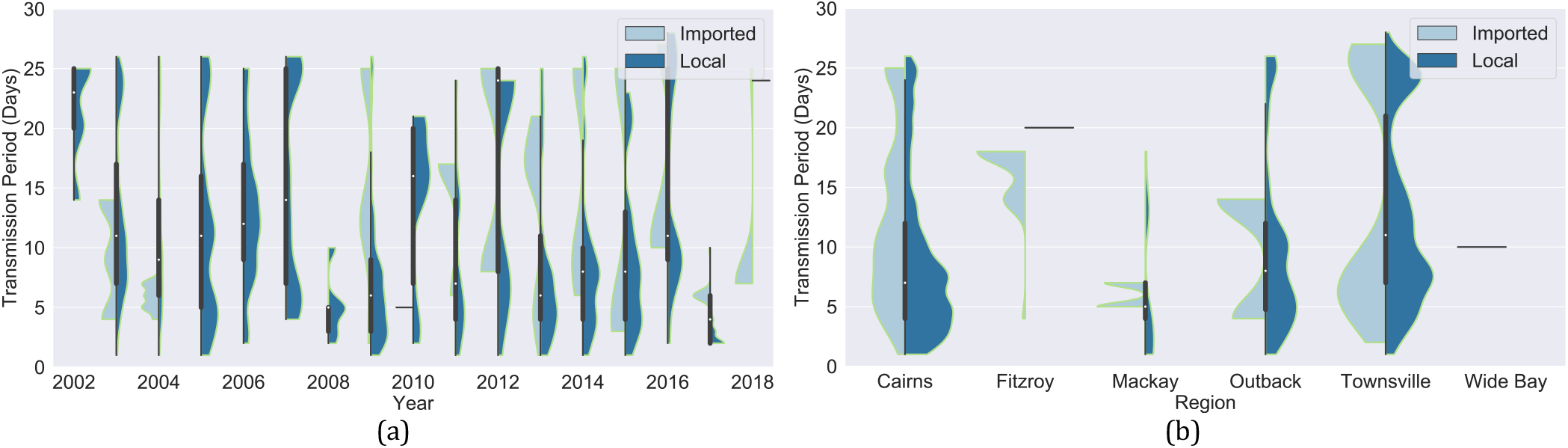
The violin charts depict the association between the fertile cases and the length of transmission period. Each element of the violin chart contains a box and whiskers plot sandwiched by the distribution curves of the transmission periods of imported and locally acquired fertile cases.

### Temporal Calibration of Response

We tested varying thresholds for what may represent efficient surveillance, and gauged their impact on various parameters associated with the incidence of dengue. In particular, we considered the impact of calibrated disease response on the occurrence and reproduction of the cases as well as the size and depth of the outbreaks. In all, we tested 6 response delay thresholds between 5 and 30 days (inclusive) with an interval of 5 days. The thresholds were enforced by removing the child cases (and their descendants) whose exposure period most likely occurred after the response efforts have been conducted for the parent case. The cases removed through this process were noted as *prevented cases*. The highest reduction was noticed across all parameters for the response delay threshold of 5 days. Another commonality was the dramatic drop in reduction between the response times of 5 and 10 days (see Figure 3).

**Figure 3.**
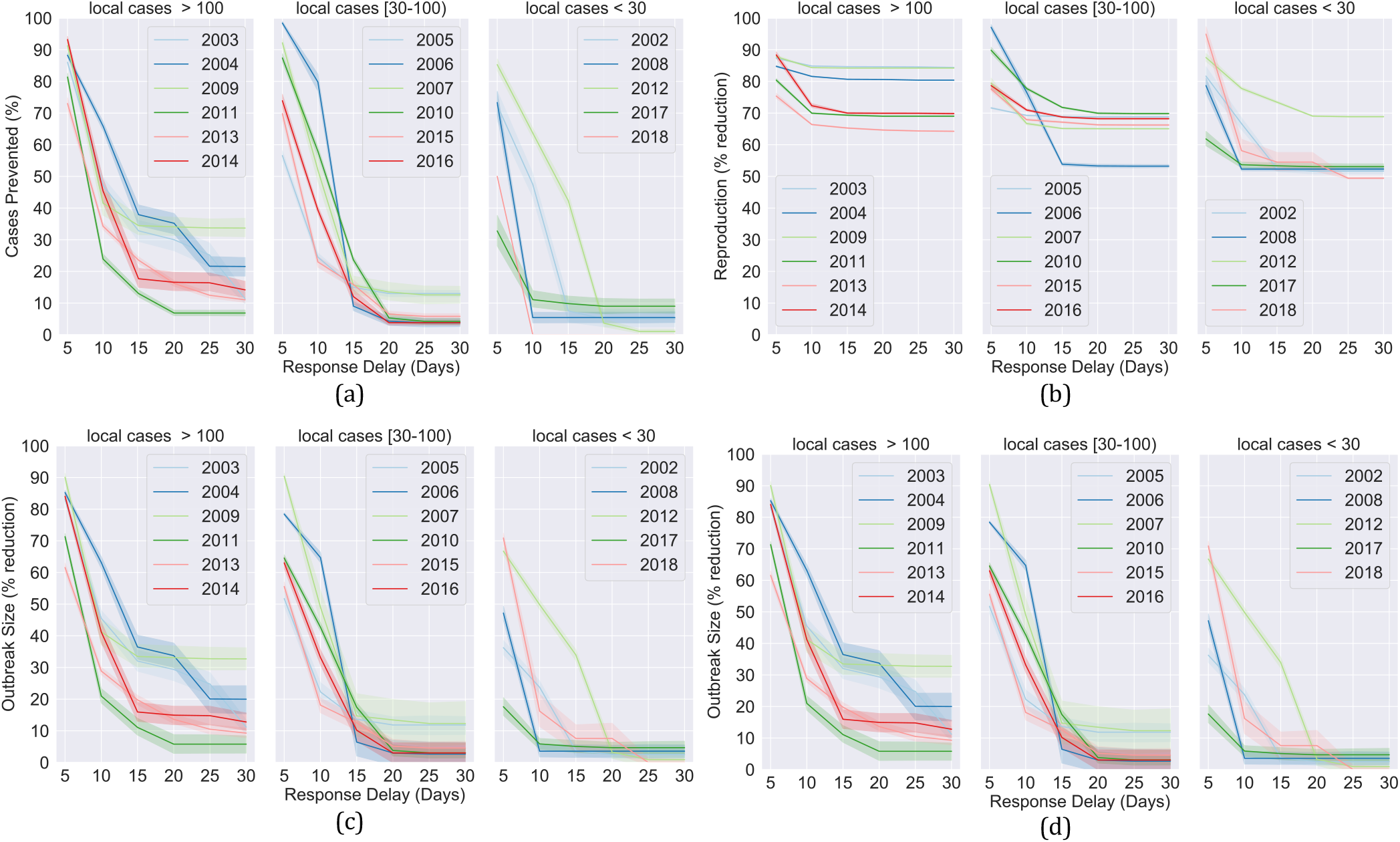
The impact of varying thresholds of response delay on dengue transmission. Figure 3a plots percentage of cases prevented while Figure 3b, c and d plot percentage reduction in reproduction, outbreak size and number of generations against varying response delay thresholds.

Notice that it is not possible to prevent a case with an unknown parent by reducing response time. Similarly, a childless case has zero reproduction and cannot contract further through reduction in response time. Therefore, we restricted our analysis of case occurrence and reproduction to the cases with a known parent and to the cases that has at least one child, respectively. We observed a large average reduction of 87% and 83% in case occurrence and reproduction respectively, when a response time of at most 5 days was enforced. A sharp decline was observed in the number of prevented cases, which contracted by 41% and a further 15% when the response delay was extended to 10 and 15 days. The difference was not as significant for higher response delay thresholds. The average reduction in case reproduction also declined with increasing response time, though not as dramatically, ranging between 71% and 83%. In terms of year-wise classification (Figure 3a), the years that recorded more than 100 locally acquired cases (especially, the top 3 years by the highest occurrence) saw a significant reduction in incidence at both ends of the response delay thresholds. On the other hand, for the years with less than 100 locally acquired cases, the proportion of prevented cases fell to less than 20% when the response delay was set to 15 days and above. The analysis of the annual breakdown of the case reproduction (Figure 3b) showed that the overall averages were skewed by the 3 years with the highest local incidence where the reduction consistently remained between 80% and 90% across all 6 response thresholds. However, this was not the case for the years with lower occurrence where a pronounced decline was noticed when the response time was extended from 5 to 10 days.

The impact of variation in response delay was also measured in terms of outbreak size and number of generations. Since an outbreak consisting of a single case cannot be reduced in size by limiting response delay, we restrict our analysis to the outbreaks of size at least 2. Similarly, when considering the reduction in number of generations, we limited our analysis to the outbreaks with at least two generations as the number of generations cannot be reduced for a singleton. Overall, the outbreak size followed a trend that was similar to that of prevented cases. An 80% [95% CI, (79, 81)] reduction was estimated in average outbreak size when the response delay was fixed at 5 days. Once again, a sharp decline (38%) was observed when the response delay was extended to 10 days with outbreaks shrinking by 42% [95% CI, (39, 45)] on average. It shrunk by another 18% (to 24% [95% CI, (21, 28)]) before flattening, when a response delay of at most 15 days was applied. The year-wise analysis of the outbreak sizes (Figure 3c) revealed trends that were quite similar to that of prevented cases. The years with less than 100 locally acquired cases showed little reduction at response thresholds of 15 days and above, while the outbreaks in years with the highest occurrence shrunk significantly at both small and large response delay thresholds. The number of generations was the least affected parameter in our analysis, with a 47% [95% CI, (45, 48)] average reduction for a response delay threshold of at most 5 days. It dropped swiftly to 19% [95% CI, (16, 21)] and then 7% [95% CI, (5, 9)] when the response delay was extended to 10 and 15 days before somewhat flattening to under 5% at the response delay of at most 25 and 30 days. Analysis of the year-wise classification (see Figure 3d) showed that a response delay of 5 days resulted in a 30 to 50% contraction in the number of generations for most years with the smallest reduction noted at 18%. This fell to 10% and below for most years when the response was delayed by 15 days. Furthermore, little to no reduction was noted at the response thresholds of 25 and 30 days.

### Reducing Response Delay

In this section we conduct a spatio-temporal analysis of the notification delay components. We divide notification delay into three parts namely, *patient delay, laboratory delay* and *clerical delay*. The patient delay is the time elapsed (in days) since the onset of symptoms to the date of specimen collection, which can be interpreted as a combination of the delay on part of a patient before they seek medical attention and the delay in clinical diagnosis by the treating doctor. The time taken by a laboratory from the date of specimen collection to the date of diagnosis is called the laboratory delay. Lastly, the time elapsed between the date of diagnosis and the date of notification is called clerical delay. A temporal plot of the notification delay and its components is provided in Figure 4a. We observe that the lab and clerical delays gradually declined as the years progressed. The average lab delay decreased from nearly 7.5 days to just over 3 days between 2003 and 2018 with minor hikes in 2006 and 2012. Similarly, the average clerical delay mostly fluctuated between 1 and 3 days for years between 2002 and 2007 (except 2004), fell to 0 in 2008 and consistently remained under 0.5 days for the following years. On the other hand, the patient delay showed no clear temporal pattern. Though a couple of dips below 3 days in the years 2008 and 2012 coincided with a drop in case fertility to under 5%, this pattern was not noticed for other year (2002, 2016, 2017 and 2018) with under 5% fertile cases. Overall, we noted that variation in notification delay from 2008 onward were largely influenced by lab and patient delays as the clerical delays flattened. Spatially (Figure 4b), Townsville saw the longest clerical delay (1.1 days) on average across all regions followed by Cairns (0.9 days), Wide Bay (0.6 days) and Queensland - Outback (0.4 days). An average lab delay of at least 4 days was reported for all regions, except Mackay. Similarly, an average patient delay of 4 days or above was observed for all regions with Townsville and Fitzroy reporting an average delay of 5 and 6 days respectively. Overall, an average notification delay of at least 9 days was noted in all regions except Wide Bay (8.1 days). Of the 3 regions with the highest incidence (Cairns, Queensland - Outback and Townsville), Townsville encountered the longest notification delay (11.1 days) on average.

**Figure 4.**
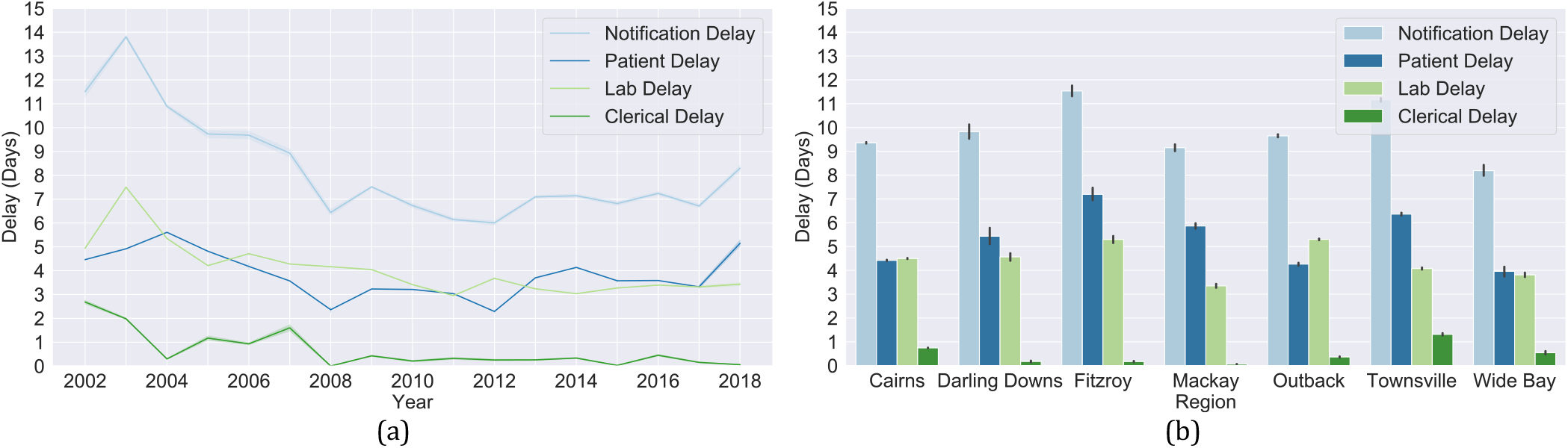
A breakdown of notification delay into three components plotted temporally Figure 4a and spatially Figure 4b.

For further insight we apply the spatial analysis of the lab and patient delays on a finer (locality) level. A finer spatial analysis can prove useful in providing guidelines on targeted resource allocation for public awareness as well as to improve diagnostic abilities and turn- around times for doctors and laboratories involved. A geo-spatial depiction of the average lab and patient delay in the localities of Cairns is provided in Figure 5. For more details on delays in each locality, box plots are also provided. Since locality names do not further the significance of our analysis and can potentially lead to re-identification, they were omitted from the box plots (x-axis). A similar spatial analysis along with an analysis of the seasonal and demographic trends of the notification delay components was also conducted for all SA4s under consideration. The results of these analyses are provided in the supplementary material.

**Figure 5.**
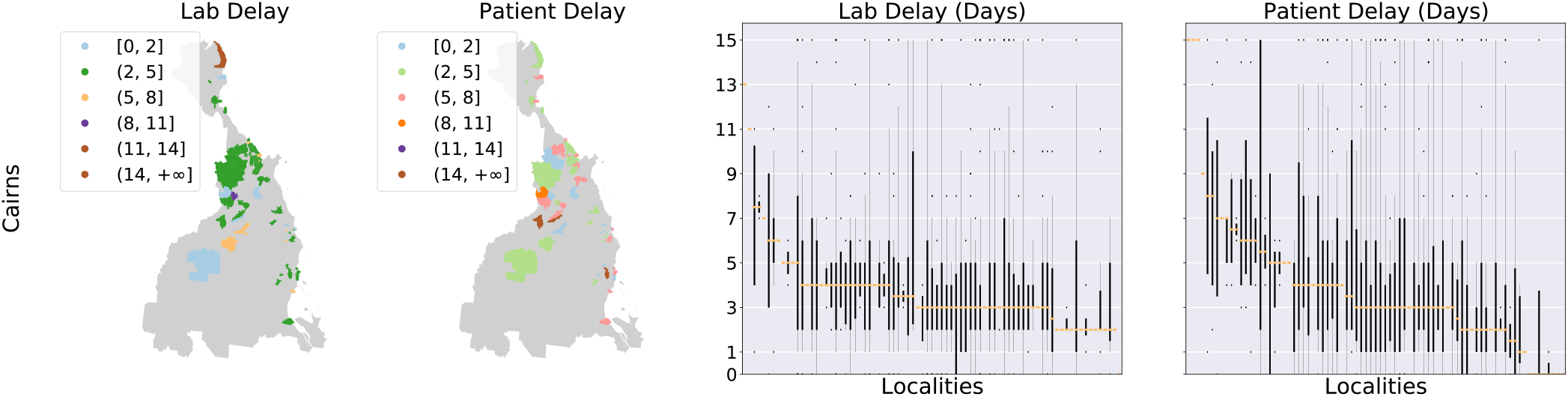
Geo-spatial depiction of the average lab and patient delays in the localities of Cairns, supplemented by the box plots.

## Discussion

We introduced the principle of epidemiological soundness that can be applied to construct a disease spread network. The construction of the network implicitly quantifies the fertility and reproduction of the cases as well as the size and depth of the outbreaks, all important parameters for an epidemiological analysis. Although our response delay threshold analysis was focused on dengue occurrence in Australia, our methodology can be applied to any infectious disease in any country.

While we confirmed that fertile cases were a result of long response delays, we found that the reverse was not true. That is, a long response delay did not necessarily lead to a fertile case. There are a range of factors that may prevent the vector from feeding on an infectious individual despite a long response delay, including housing with air conditioning and fly nets, the use of mosquito repellent and confinement to a mosquito free space (e.g., hospital) during illness. Moreover, vector presence data is collected through sampling (by setting up mosquito traps at various locations in a region). While this may be the only viable option, it is insufficient to confirm the presence of the vector in all parts of a region. The involvement of these factors and a lack of data on them prevent us from analysing the relationship between response delay and infertile cases.

Most disease surveillance systems are affected by a degree of under-ascertainment leading to uncertainty on the actual incidence of the disease. Our analysis highlighted the under-ascertainment of dengue occurrence in Queensland. Given that dengue is non-endemic in Australia, each outbreak is expected to be rooted in an overseas acquired case. However, while constructing the disease spread network, we were unable to identify a parent case for approximately 16% of the locally acquired cases. Furthermore, the years 2002 and 2005 to 2008 were estimated to have no imported fertile cases despite having local transmission. All indicative of the under-ascertainment of dengue occurrence in Australia.

A short EIP and delayed response have been previously attributed to be the major contributing factors for large dengue outbreaks in 2009 [24]. Our analysis confirmed this and revealed that it was also the case for other years of large dengue outbreaks including 2003, 2004 and 2014, by noticing significant reduction in average occurrence, reproduction and outbreak size at both small and large response delay thresholds.

A sharp decline was observed in reduction of outbreak generations at the response thresholds of 10 and 15 days indicating that in most instances the outbreaks have a smaller inter-generation gap. This observation is a possible explanation for a similar fall in the reduction of occurrence and outbreak size as the response efforts may be committed when the first few (one and possibly more) generations of cases have already produced child cases.

The Queensland Health Dengue Management Plan states that the treating doctors are required to report any suspicious cases of dengue before the laboratory confirmed diagnosis. However, over the 16 years of dengue occurrence covered in this study we found only 34 (0.6%) cases that were notified before a laboratory diagnosis, stressing the need for a wider awareness about the importance of reporting suspicious cases and better understanding of the dengue symptoms for clinical diagnosis.

We observed that the number of locally acquired cases in Cairns and Townsville regions remained consistently low over the last few years of the study (14 and 11 cases respectively since 2016) despite longer than ideal response delays. We note that this unusual trend can be attributed to the deployment of the Wolbachia infected mosquitoes in these regions since 2011 [25].

Our work points to various future research directions. One interesting extension of the study would be the development of a case ranking system for laboratories, where cases would be ranked by a risk function. The risk function could be based on various parameters including time since onset, place of residence and weather conditions at the place of residence since onset. Another interesting problem to pursue would be to estimate the economic burden of inefficient surveillance. It could cover among other costs, the spending on providing healthcare for larger number of patients as the outbreaks grow and the economic losses due to days spent off work.

## Conclusion

We introduce the principle of epidemiological soundness, which utilizes a combination of the disease occurrence, meteorological and human population mobility data, to construct a disease spread network. Using the network, we empirically confirm that high morbidity relates positively with delay in disease response. Moreover, we identify what constitutes efficient surveillance by applying various thresholds of the disease response delay to the network and report their impact on case fertility, reproduction, outbreak size and number of generations. Lastly, we identify the components of the disease surveillance system that can be calibrated to achieve the identified efficiency thresholds. Our methodology may be of independent interest, irrespective of the underlying data and can be utilized to provide guidelines on spatially and demographically targeted resource allocation for public awareness campaigns as well as to improve diagnostic abilities and turn-around times for the doctors and laboratories wherever required. The surveillance efficiency thresholds obtained in this study can be utilized to define a case ranking system for the pathology labs that can help prioritize testing of the cases that are more likely to lead to a larger outbreak.

## Data Availability

The datasets underlying the results of this study are available at the discretion of the data custodians due to associated legal, ethical and privacy concerns. The access to dengue occurrence data is restricted by Queensland Health in accordance with the Public Health Act 2005. Interested parties can request access through the Communicable Diseases Branch of Queensland Health. The IVS and NVS data can be obtained from Tourism Research Australia. Weather data can be obtained upon request from Bureau of Meteorology (http://www.bom.gov.au/climate/data-services/data-requests.shtml). All the source code necessary to replicate the presented results is available upon request on a case-to-case basis due to commercial considerations.

## Declarations

### Ethics Approval

The study was approved by the CSIRO Social Science Human Research Ethics Committee (Ethics Clearance 065/19) and by the Health Innovation, Investment and Research Office (HIIRO) under section 284 of the Public Health Act 2005 (grant number: RD007950).

## Acknowledgements

We acknowledge the contributions of Communicable Diseases Branch of Queensland Health, the Australian Bureau of Meteorology and Tourism Research Australia for providing data used in this study. We would like to thank Frank de Hoog and Roslyn Hickson for their valuable comments, which greatly helped to improve the manuscript.

