## supplementart material for "Efficient Surveillance and Temporal Calibration of Disease Response"

### FINER ANALYSIS OF THE NOTIFICATION DELAY COMPONENTS

We conducted further analyses to identify regional and demographic classes of the patients, doctors and labs that need to be the focus of targeted campaigns to avoid longer patient and lab delays. In order to evaluate the impact of seasonality on the performance of the labs, we classified the reported dengue cases by their month of onset. Figure S 1a depicts the seasonal trends of dengue occurrence. We noticed that lab delays peaked during the 4th quarter of the year with more than 50% of cases encountering a lab delay of at least 4 days, while the smallest delays occurred in the 3rd quarter. The 3rd and 4th quarter coincide with the off-peak and peak season of dengue occurrence in Queensland, respectively.  One interpretation of this trend is that the performance of the labs drops under higher case load which may be due to limited testing capacity. However, this interpretation is countered to an extent by a decline in lab delays recorded in January and February, the months that make up the later part of the peak dengue season in Queensland. We speculate that this may be attributed to the added vigilance of the laboratory staff due to the occurrence of historic outbreaks during this part of the year. To gain further insight we break down seasonal lab delays by SA4s (see Figure S 1b). In this context, we noticed a jump in average lab delays for cases reported during April, October and December in the Cairns region. Similarly, for cases reported in Townsville, the delays peaked during September and October.  Interestingly, April and October are also the months of school holidays in Queensland, which leads to the possibility that the longer lab delays during these months may be due to the diminished capacity of the labs with staff taking time off work. We were unable to identify any meaningful patterns for the rest of the regions. We believe that the lack of a clear trend may in part be due to the limited number of cases recorded in these regions.

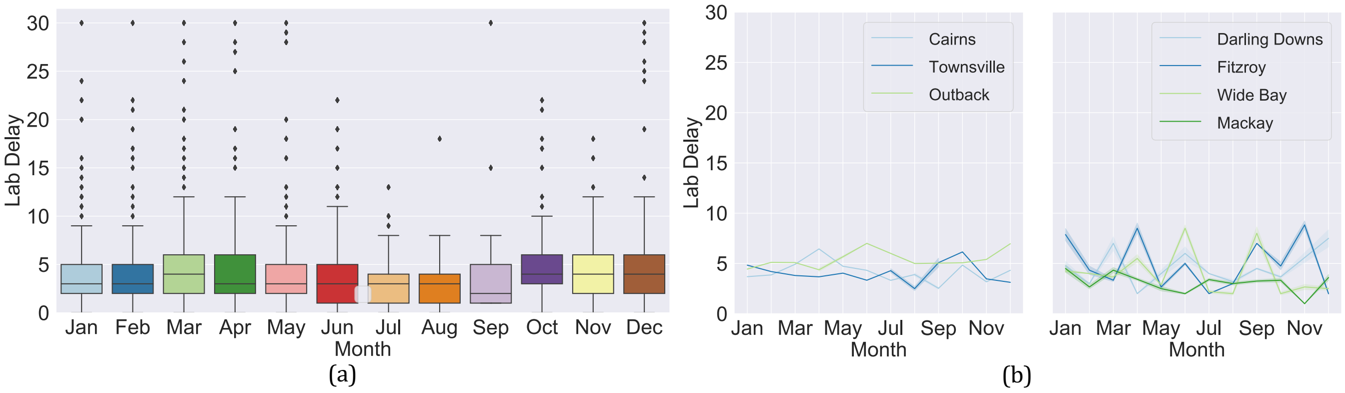


*Figure S 1. The overall and region-wise seasonal trends of lab delay. The region-wise trends are split into two subgraphs to improve visibility.*

We also evaluated the impact of seasonality on the patient delays (see Figure S 2a). We observed that the patient delays peaked during the spring season between August and October. Since the spring season is also the peak time for hay fever in Queensland, which presents symptoms that are similar to that of dengue, the rise is patient delays may be due to misattribution of the dengue symptoms to hay fever.

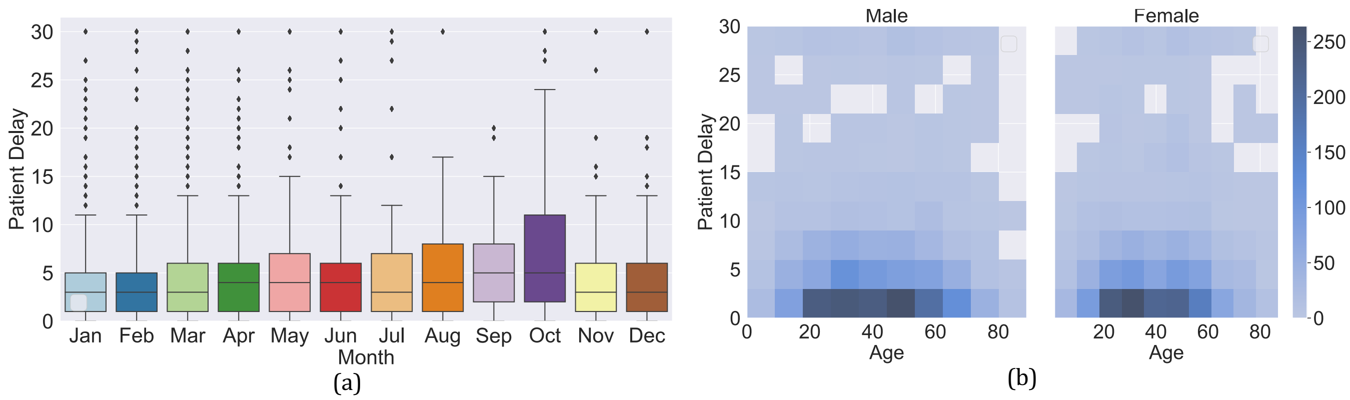


*Figure S 2. The seasonal and demographic classification of the patient delay.*

Furthermore, in order to identify demographic groups for targeted awareness campaigns, we performed an analysis of the patient delay trends through classification of the reported cases by age and gender (see Figure S 2b). We found that in male patients, most cases with a patient delay of 4 to 5 days were observed in the age group 30 to 35 years. Moreover, the largest number of cases with a delay of 10 or more days were observed in patients of age 45 to 60. On the other hand, a delay of 4 to 5 days was most frequently observed in female patient of age groups 20-35 and 45-50 years.

Lastly, we present the extended results of the finer spatial analysis briefly described in the main text. A geo-spatial depiction of the average lab and patient delay in the localities of the SA4s considered in this study (except Cairns) are provided in **Error! Reference source not found.**. For more details on delays in each locality, box plots are also provided. The results presented here can prove useful in providing guidelines on development of spatially targeted campaigns and resource allocation for public awareness as well as to improve diagnostic abilities and turn-around times for the doctors and laboratories involved.

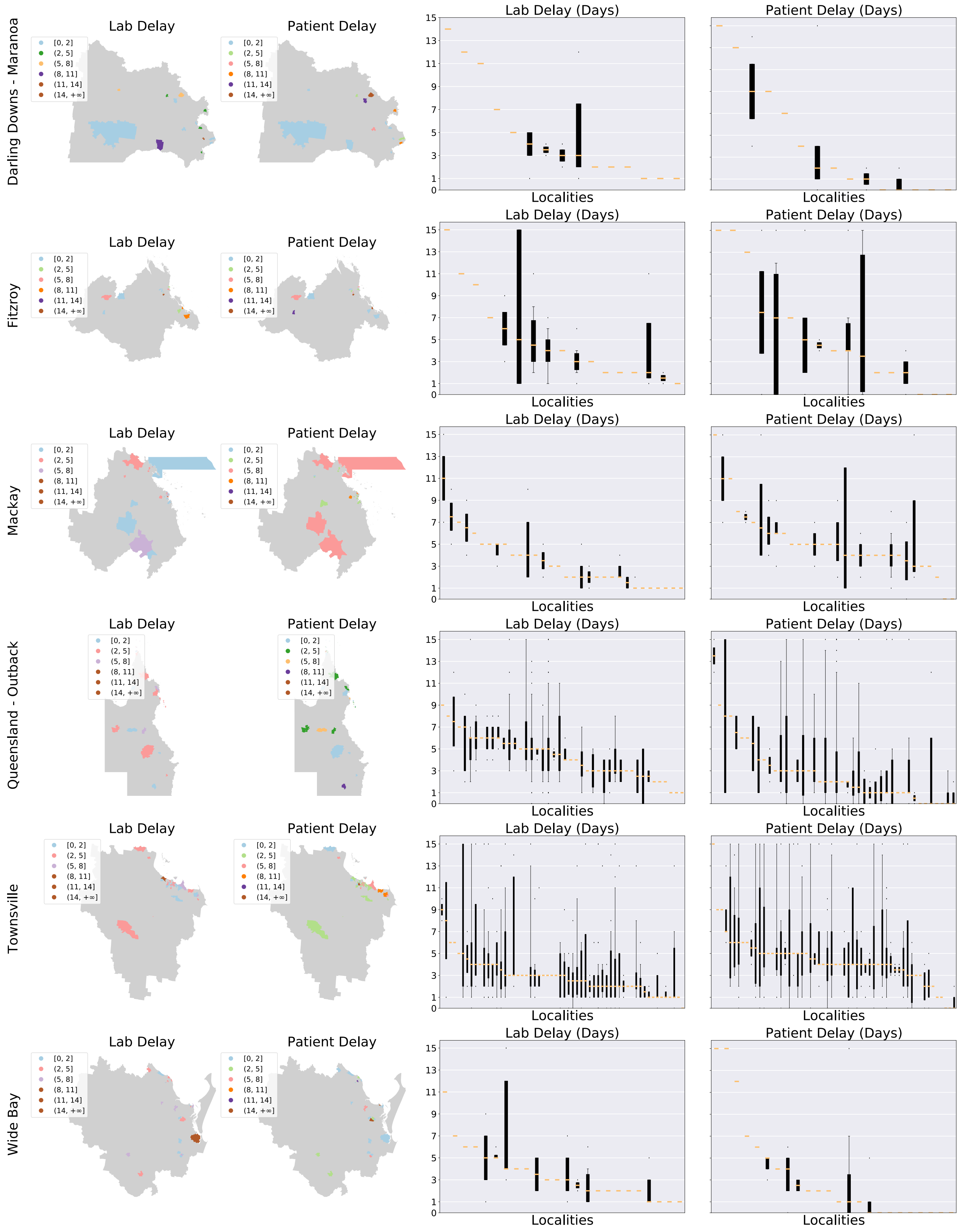


*Figure S 3. Geo-spatial depiction of the average lab and patient delays in the localities of the vector present regions (excluding Cairns), supplemented by the box plots.*
